# Early Life Predictors of Obesity and Hypertension Comorbidity at Midlife: Findings from the 1958 National Child Development Study (NCDS)

**DOI:** 10.1101/2024.12.09.24318705

**Authors:** S Stannard, RK Owen, A Berrington, N Ziauddeen, SDS Fraser, S Paranjothy, RB Hoyle, N A Alwan

**Author notes:** Corresponding Author: Sebastian Stannard -, University of Southampton, University Road, Southampton, SO17 1BJ, United Kingdom.

## Abstract

**Background:** Early life exposures can increase the risk of both obesity and hypertension in adulthood. In this paper we identify exposures across five pre-hypothesised childhood domains, explore them as predictors of obesity and hypertension comorbidity using the 1958 National Child Development Study (NCDS), and discuss these results in comparison to a similar approach using another birth cohort (the 1970 British Cohort Study (BCS70)).

**Methods:** The analytical sample included 9150 participants. The outcome was obesity (BMI of ≥30) and hypertension (blood pressure>140/90mm Hg) comorbidity at age 44. Domains included: ‘prenatal, antenatal, neonatal and birth’, ‘developmental attributes and behaviour’, ‘child education and academic ability’, ‘socioeconomic factors’ and ‘parental and family environment’. Stepwise backward elimination selected variables for inclusion for each domain, and predicted risk scores of obesity-hypertension for each cohort member within each domain were calculated. We performed multivariable logistic regression analysis including domain-specific risk scores, sex and ethnicity to assess how well the outcome could be predicted taking all domains into account. In additional analysis we included potential adult factors.

**Results:** Including all domain-specific risk scores, sex, and ethnicity in the same prediction model the area under the curve was 0.70 (95%CI 0.67-0.72). The strongest domain predictor for obesity-hypertension comorbidity was for the socioeconomic factors domain (OR 1.28 95%CI 1.18-1.38), similar to the BCS70 results. However, the parental and family environment domain was not a significant predictor for obesity-hypertension comorbidity (OR 1.08 95%CI 0.94-1.24) unlike the BCS70 results. After considering adult predictors, robust associations remained to the socioeconomic, education and academic abilities, development and behaviour, and prenatal, antenatal, neonatal and birth domains.

**Conclusions:** In the NCDS some early life course domains were found to be significant predictors of obesity-hypertension comorbidity, supporting previous findings. Shared early-life characteristics could have a role in predicting obesity-hypertension comorbidity, particularly for those who faced socioeconomic disadvantage.

## Introduction

In England an estimated 26% of adults have obesity (body mass index 30kg/m^2^ or over) [1] and 31% of adults have hypertension (blood pressure over 140/90mm Hg) [2]. Both conditions are associated with further morbidities including Type 2 diabetes, heart disease, kidney disease, renal disease, strokes, and some cancers, including breast and bowel cancer [3–6]. Globally, both conditions represent a major public health problem. The World Health Organisation (WHO) estimates that 2.8 million deaths every year are attributed to overweight or obesity [7]. Hypertension prevalence has increased between 41-144% across WHO regions over the past 30 years [8]. Research has also demonstrated that obesity and hypertension often co-occur [9,10]. For example, around half of hypertensive patients in the US have obesity [11], and individuals with obesity are 3.5 times more likely to have hypertension compared to normal weight individuals [12,13]. The co-occurrence of both conditions also increases the risk of further morbidity including cardiovascular disease, sexual dysfunction, poor quality of life and mortality [13,14].

It is important to consider risk exposures to subsequent morbidity in terms of multiple domains (i.e., a group of variables that represent an overarching theme) for three reasons. First, this provides a combined exposure measure that reflects multiple variables in the data rather than performing multiple statistical testing using single components in relation to the study outcomes. Second, conceptualising the components within wider early life domains provides a more holistic reflection of the childhood conditions in which the cohort member grew up. Third, considering early life risk in domains may better inform interventions and policy in childhood as incorporating information from multiple early life domains into the same analysis may help us understand the combined effects of different experiences across a range of early life domains on developing long-term conditions. This can provide actionable insights into developing interventions in childhood that may help support people to live longer in good health.

In previous research [15], we developed a conceptual framework to characterise the population-level domains of early-life determinants of future multimorbidity risk. Through a scoping literature and policy review, and with the support of public and patient contributors, 12 domains of early-life risk factors of future multimorbidity risk were identified [15]. These domains covered a range of social, economic, developmental, educational and environmental factors that focused on both direct and indirect factors as well as wider systemic and structural determinants of disease and health inequalities [15,16]. Subsequent research [17] explored how these 12 domains of early-life risk factors of future multimorbidity risk could be characterised across UK birth cohort studies including the 1958 National Child Development Study (NCDS).

In a previous paper we used the 1970 British Cohort Study (BCS70) to consider five early life domains, chosen because they showed unadjusted associations with obesity and hypertension comorbidity at age 46 [18]. Results indicated all five domains were significant predictors for obesity-hypertension comorbidity, with the strongest domain predictors being the parental and family environment domain and the socioeconomic factors domain [18]. After including adult predictors, the most robust domains for predicting obesity-hypertension included the parental and family environment domain, socioeconomic factors domain and education and academic ability [18].

In this analysis, we aimed to apply the same methodological approach to the NCDS cohort. Although the two cohorts are only 12 years apart, the childhood conditions the members of each cohort grew up in are arguably different. The younger cohort (BCS70) experienced rising women’s employment and economic independence, decreased family stability, educational expansion and a shift away from the male breadwinner nuclear family that dominated the family structure of the NCDS cohort [19–23].

The younger cohort (BCS70) also experienced generational pay progression, higher real household disposable incomes and an increase in home ownerships [19], but globalisation and a shift of income from labour to capital meant greater economic uncertainty and increased inequality [23]. Therefore, these factors might lead us to expect more heterogeneity in the early life experiences of the 1970 cohort compared to the 1958 cohort.

In this study, we aimed to explore the extent to which the same five early life course domains predict the outcome of obesity-hypertension comorbidity. To achieve this while weighting the components of each domain, we also produced-as a first stage of the analysis-predicted risk scores of the outcome for each of the five pre-defined early life domains. In the discussions we consider the results presented here in relation to a previous analysis of the BCS70 cohort [18]. This work forms part of a larger aim to model targeted multimorbidity prevention scenarios as part of the Multidisciplinary Ecosystem to study Lifecourse Determinants and Prevention of Early-onset Burdensome Multimorbidity (MELD-B) project [24].

## Methods

### Datasets

We used the 1958 National Child Development Study (NCDS) [25] that has followed children born in England, Scotland and Wales in one week in 1958, and includes 17415 cohort members. The comorbidity outcome of obesity and hypertension was measured in a biomedical sweep at age 44. All other variables were measured either at birth or at the age 11 sweep.

### Outcome

The outcome was a combined obesity-hypertension phenotype at age 44. Blood pressure was measured via three systolic and diastolic blood pressure readings during a single appointment from a research nurse. Hypertension was defined as average blood pressure reading of over 140/90 mm Hg. BMI was calculated via height and weight measurements taken during the same nurse appointment, and calculated using the following formula: BMI = weight (kg) / height (m)^2^. Obesity was defined as a BMI of 30kg/m^2^ or over. The obesity-hypertension comorbidity variable was considered as a binary (no/yes) variable.

### Exposures (Five pre-hypothesised domains)

We focus on five domains because they showed unadjusted association with the outcome in this paper and in the previous research based on the BCS70 cohort [18], and Supplementary Materials Table 1 includes all the variables that were considered for each domain:

1. *Prenatal, antenatal, neonatal and birth domain* focused on the period from conception to the onset of labour, the circumstances and outcomes surrounding a birth, and the period immediately following birth.
2. *Developmental attributes and behaviour domain* focused on the developmental markers of children relating to cognition, coordination, personality types and behavioural traits.
3. *Child education and academic ability domain* related to the process of learning and educational achievement, especially in educational settings, and the knowledge an individual gains from these educational institutions.
4. *Socioeconomic factors domain* included factors relating to differences between individuals or groups of people caused mainly by their social and economic situation.
5. *Parental and family environment domain* incorporated the interactions between children and care givers, parenting styles, parental beliefs, attitudes and discipline, and wider family factors such as kin networks.

**Table 1.**
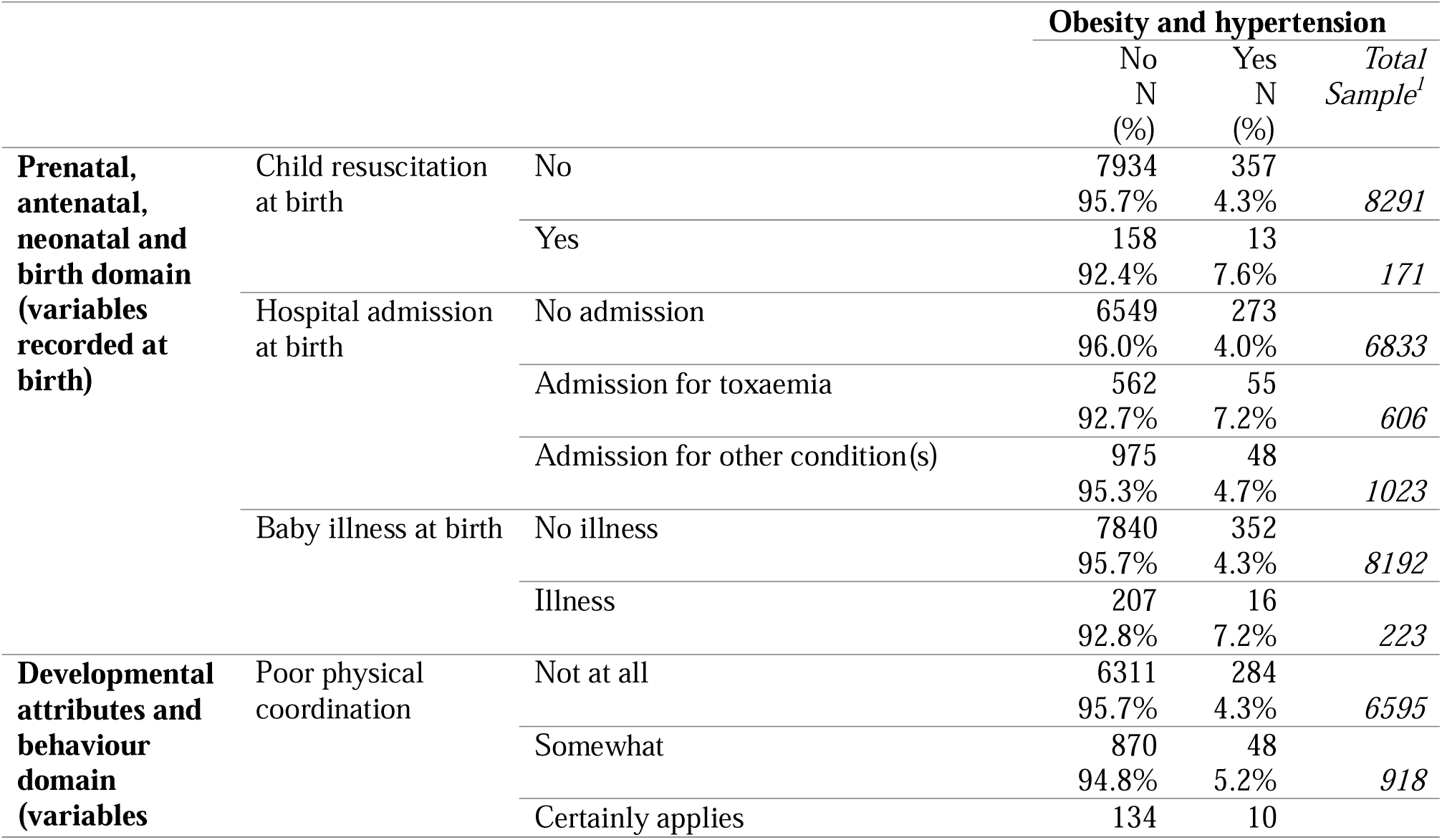
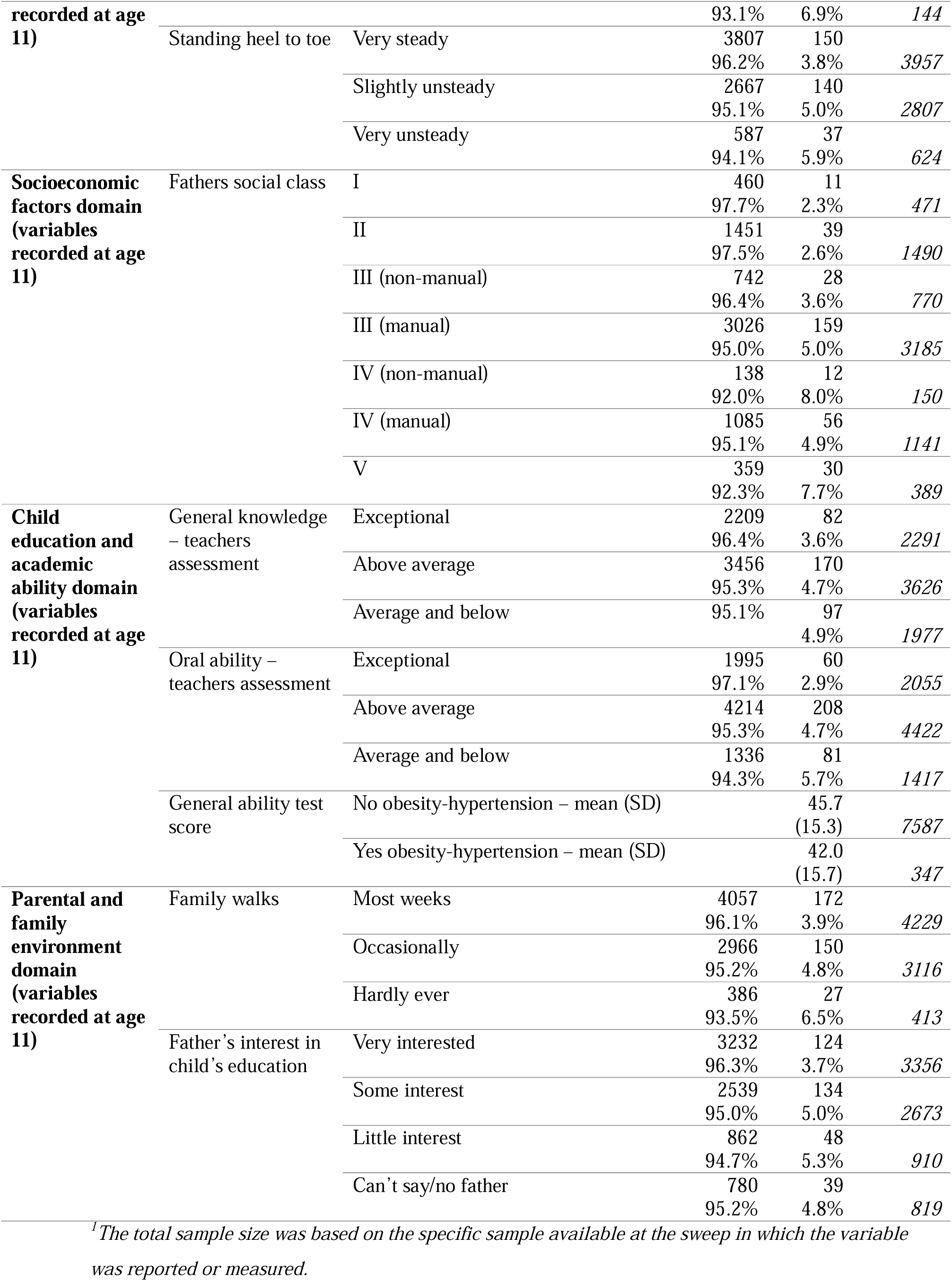
Step 1: Variables retained in the five domain risk scores following stepwise backwards elimination, and prevalence of combined obesity-hypertension at age 44.

### Analytical sample

The analytical sample included all cohort members who had measured height, weight, and blood pressure at age 44 (n = 9150/17415). Multiple imputation was used to maintain sample size and reduce bias in the estimates due to missing data. Multiple imputation was conducted by chained equations for missing observations at birth, age 11, and 44 [26]. 50 imputation cycles were constructed under the missing-at-random assumption [27–29], which has been found to be highly plausible in the British birth cohorts [30]. All variables were included in the imputation process. The outcome was included in the imputed models, but imputed outcome values were not used.

### Statistical analysis

We used the same methodological approach followed to examine five early-life domains as predictors of obesity-hypertension comorbidity in the BCS70 cohort, and full details of this method can be found elsewhere [18]. However, we provide a summary of those methods here.

Firstly, stepwise backward elimination conducted on multiple imputed data, was used to select variables for inclusion separately for each domain. Secondly, logistic regression models then explored the relationship between retained variables following stepwise backwards elimination and odds of obesity-hypertension comorbidity. From this regression modelling the predicted risk scores of the obesity-hypertension comorbidity outcome for each cohort member within each domain were then calculated. Thirdly, we used logistic regression modelling including all five domain-specific risk scores, and added sex (determined at birth by the presence or absence of a Y chromosome), and ethnicity to predict the obesity-hypertension comorbidity outcome. The area under curve statistic was used to assess the predictive performance of this model. The odds ratios of the five domains within this model were used to identify the strongest domains that acted as predictors for obesity and hypertension comorbidity taking into account the effect of the other domains. Finally, we repeated the previous step this time with the inclusion of adult factors that are potentially linked to both the exposures and the outcome. These were recorded at age 44 and included marital status, self-rated financial difficulty, the age the cohort member left education, smoking status, frequency of consuming alcoholic drinks, frequency of consuming fresh fruit, frequency of exercising and occupational social class.

### Ethical Approval

Ethics approval for MELD-B was obtained from the University of Southampton Faculty of Medicine Ethics committee (ERGO II Reference 66810). A full review of ethics and consent for the NCDS study is provided by the Centre for Longitudinal Studies and can be found elsewhere [31].

## Results

### Step 1: Stepwise backwards elimination to select variables for inclusion separately for each domain

Table 1 identifies the retained variables following stepwise backwards elimination for each domain in relation to the outcome of obesity-hypertension comorbidity at age 44. Amongst the 9150 cohort members at age 44, 4% (n=395) had obesity-hypertension comorbidity. Considering the individual prevalence, 24% of the cohort members at age 44 had obesity (n=2235/9150) and 11% had hypertension (n=1049/9150). For the prenatal, antenatal, neonatal and birth domain, the retained variables following stepwise backwards elimination focused on the health of the cohort member at birth (illness, resuscitation and admission to hospital). For the developmental attributes and behaviour domain, retained variables considered developmental markers of the cohort members (coordination and balance), whilst father’s social class was retained within the socioeconomic factors domain. For the child education and academic ability domain, retained variables were related to academic ability (general ability test, general knowledge assessment and oral ability assessment), and for the parental and family environment domain, retained variables focused on fathers’ interest in their child’s education and engaging in family activities (family walks).

### Step 2: Predicted risk scores of obesity-hypertension comorbidity for each cohort member within each domain

In Supplementary Materials Table 3-7, we include the regression coefficients of obesity-hypertension for the logistic regression models that explored the relationship between retained variables following stepwise backwards elimination (step 1) and obesity-hypertension comorbidity, and for each domain separately. In Supplementary Materials 2, we present a Pearson correlation matrix exploring correlation across domains. The strongest correlations were between the parental and family environment domain and child education and academic ability domain (coefficient 0.31) and between the parental and family environment domain and socioeconomic factors domain (coefficient 0.29). These results suggest that the domain-specific risk scores were not collinear.

### Step 3: A prediction model including five domain-specific risk scores, sex and ethnicity

The area under curve (AUC) for the prediction model that included all five domain-specific risk scores (produced in step 2), ethnicity, and sex was 0.70 (95%CI 0.67-0.72). Table 2 presents the odds ratios of obesity-hypertension at age 44 for the prediction model that included all five domain-specific risk scores (produced in step 2), sex, and ethnicity. As shown, the parental and family environment domain was not a significant predictor of obesity-hypertension comorbidity (OR 1.08 95%CI 0.94-1.24). All other domains were predictors of obesity-hypertension comorbidity, with the strongest domain predictor being the socioeconomic factors domain (OR 1.28 95%CI 1.19-1.38).

**Table 2.** Step 3: Odds ratios of obesity-hypertension at age 46 in relation to domain-specific risk score of obesity-hypertension for five early life domains. Multiple imputed data (50 Imputations).

|  | Odds<br>ratios | 95% Confidence<br>interval |
| --- | --- | --- |
| Developmental attributes and behaviour domain | **1.20** | **1.06 - 1.36** |
| Prenatal, antenatal, neonatal and birth domain | **1.21** | **1.11 - 1.31** |
| Parent and family environment domain | 1.08 | 0.94 - 1.24 |
| Education and academic ability domain | **1.12** | **1.02 - 1.23** |
| Socioeconomic factors domain | **1.28** | **1.19 - 1.38** |
| Child's sex: Women (base – men) | **0.38** | **0.64 - 0.91** |
| Mother ethnic group: Other (base – white) | 1.37 | 0.86 - 2.21 |
| Number of observations |  | 9150 |

*Significant findings highlighted in bold.*

**Table 4.**
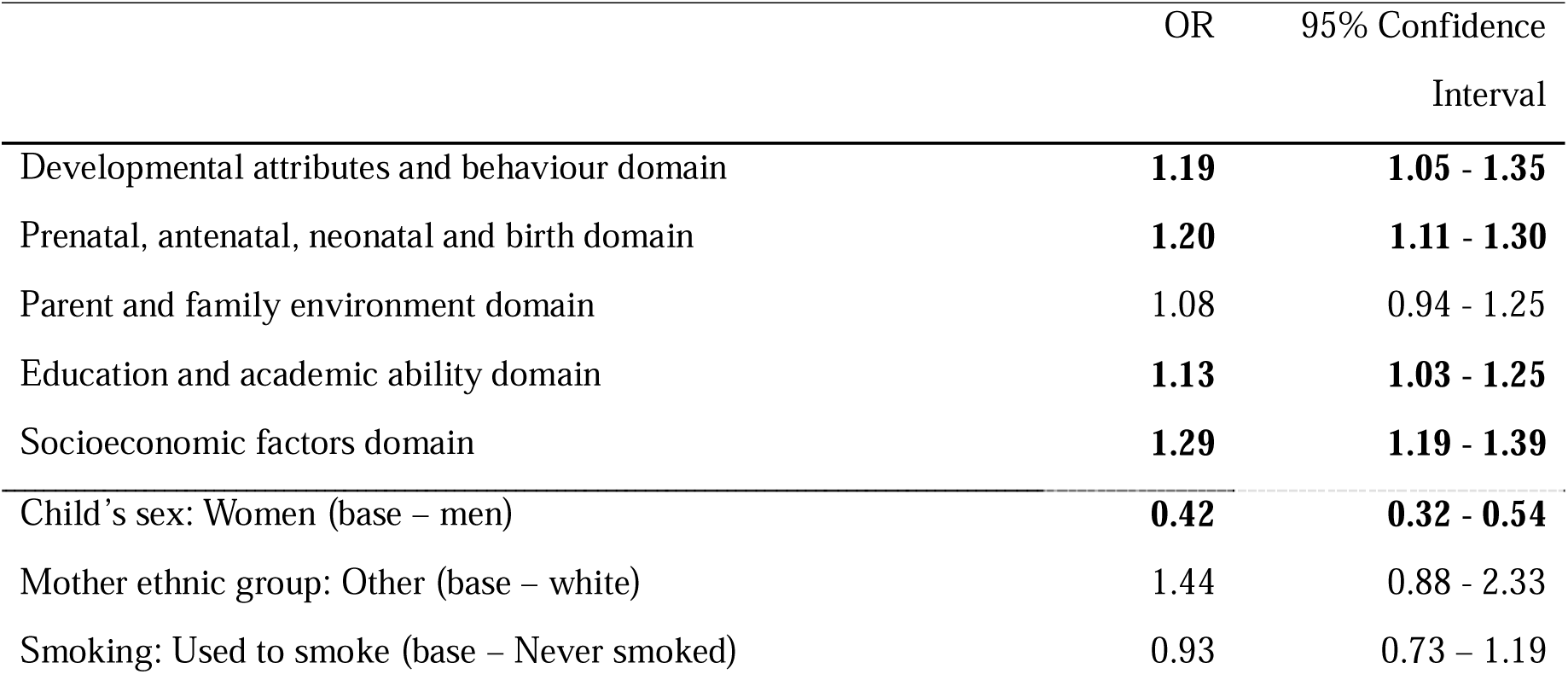
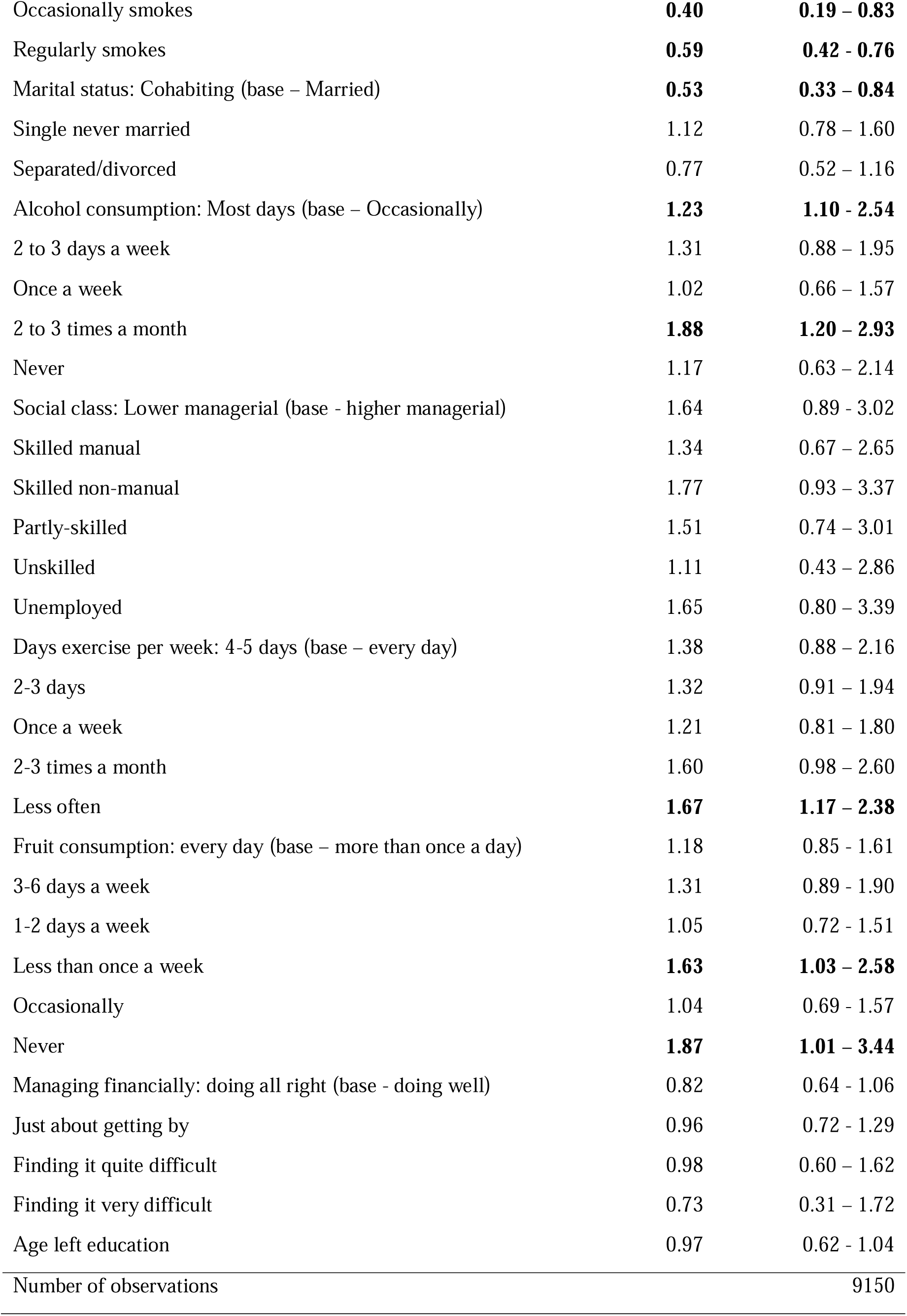
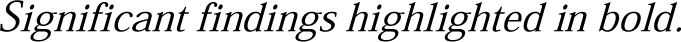
Step 4: Odds ratios of obesity-hypertension at age 46 in relation to domain-specific risk score of obesity-hypertension for five early life domains including adult predictors. Multiple imputed data (50 Imputations).

|  | OR | 95% Confidence Interval |
| --- | --- | --- |
| Developmental attributes and behaviour domain | **1.19** | **1.05 - 1.35** |
| Prenatal, antenatal, neonatal and birth domain | **1.20** | **1.11 - 1.30** |
| Parent and family environment domain | 1.08 | 0.94 - 1.25 |
| Education and academic ability domain | **1.13** | **1.03 - 1.25** |
| Socioeconomic factors domain | **1.29** | **1.19 - 1.39** |
| Child's sex: Women (base – men) | **0.42** | **0.32 - 0.54** |
| Mother ethnic group: Other (base – white) | 1.44 | 0.88 - 2.33 |
| Smoking: Used to smoke (base – Never smoked) | 0.93 | 0.73 – 1.19 |
| Occasionally smokes | **0.40** | **0.19 – 0.83** |
| Regularly smokes | **0.59** | **0.42 - 0.76** |
| Marital status: Cohabiting (base – Married) | **0.53** | **0.33 – 0.84** |
| Single never married | 1.12 | 0.78 – 1.60 |
| Separated/divorced | 0.77 | 0.52 – 1.16 |
| Alcohol consumption: Most days (base – Occasionally) | **1.23** | **1.10 - 2.54** |
| 2 to 3 days a week | 1.31 | 0.88 – 1.95 |
| Once a week | 1.02 | 0.66 – 1.57 |
| 2 to 3 times a month | **1.88** | **1.20 – 2.93** |
| Never | 1.17 | 0.63 – 2.14 |
| Social class: Lower managerial (base - higher managerial) | 1.64 | 0.89 - 3.02 |
| Skilled manual | 1.34 | 0.67 – 2.65 |
| Skilled non-manual | 1.77 | 0.93 – 3.37 |
| Partly-skilled | 1.51 | 0.74 – 3.01 |
| Unskilled | 1.11 | 0.43 – 2.86 |
| Unemployed | 1.65 | 0.80 – 3.39 |
| Days exercise per week: 4-5 days (base – every day) | 1.38 | 0.88 – 2.16 |
| 2-3 days | 1.32 | 0.91 – 1.94 |
| Once a week | 1.21 | 0.81 – 1.80 |
| 2-3 times a month | 1.60 | 0.98 – 2.60 |
| Less often | **1.67** | **1.17 – 2.38** |
| Fruit consumption: every day (base – more than once a day) | 1.18 | 0.85 - 1.61 |
| 3-6 days a week | 1.31 | 0.89 - 1.90 |
| 1-2 days a week | 1.05 | 0.72 - 1.51 |
| Less than once a week | **1.63** | **1.03 – 2.58** |
| Occasionally | 1.04 | 0.69 - 1.57 |
| Never | **1.87** | **1.01 – 3.44** |
| Managing financially: doing all right (base - doing well) | 0.82 | 0.64 - 1.06 |
| Just about getting by | 0.96 | 0.72 - 1.29 |
| Finding it quite difficult | 0.98 | 0.60 – 1.62 |
| Finding it very difficult | 0.73 | 0.31 – 1.72 |
| Age left education | 0.97 | 0.62 - 1.04 |
| Number of observations |  | 9150 |

*Significant findings highlighted in bold.*

### Step 4: A prediction model including five domain-specific risk scores, sex, ethnicity and potential adult predictors

Including adult factors in the model, the AUC increased to 0.73 (95%CI 0.71-0.76). Table 3 presents the odds ratios of obesity-hypertension at age 44 for this prediction model that included all five domain specific risk scores (produced in step 2) ethnicity, sex and adult factors. As shown, after the inclusion of adult factors, the prenatal, antenatal, neonatal and birth domain was not a significant predictor of obesity-hypertension comorbidity (OR 1.08 95%CI 0.94-1.25). All the other four domains remained robust predictors for obesity and hypertension comorbidity, with the strongest domain predictor for obesity and hypertension comorbidity remaining as the socioeconomic factors domain (OR 1.29 95%CI 1.19-1.39).

## Discussion

Using the NCDS birth cohort dataset, and considering five early life domains concurrently, the strongest domain predictor for obesity-hypertension comorbidity was the socioeconomic factors domain, even after accounting for socioeconomic factors during adulthood. Results suggested that the parental and family environment domain was not a significant predictor for obesity-hypertension comorbidity in adulthood. The inclusion of adult factors slightly improved the predictive performance of the models, however robust associations remained to the development and behaviour, the socioeconomic factors, education and academic abilities and prenatal, antenatal, neonatal and birth domains.

Our aim in this paper was to use the same methodological approach that has already been applied to the BCS70 cohort [18] to explore the relationship between five early life domains and obesity-hypertension comorbidity, and then compare results across the two cohort studies. We found that the results presented here were largely comparable to our previous findings based on the BCS70 cohort [18], in particular there was consistency with regards to the robustness of the socioeconomic factors domain as a predictor for obesity-hypertension comorbidity across both papers [18]. Significantly, the socioeconomic factors domain (in childhood) remained an important predictor even after accounting for socioeconomic factors during adulthood. This adds to a body of literature that has found childhood socioeconomic factors to be associated to multimorbidity later in the life course [32–36], and provides further justification for interventions targeting socioeconomic conditions in childhood as this might have the biggest impact on improving certain health outcomes later in the life course. The findings that the socioeconomic domain remains important across cohorts is significant given the BCS70 cohort (born 1970) were exposed to better economic conditions including higher employment rates particularly for women, generational pay progression, higher real household disposable incomes and an increase in home ownerships [19].

Overall, the AUC statistics suggested that for both models (including and excluding adult predictors), variables within the NCDS cohort had a stronger predictive power than variables within the BCS70 cohort. Odds ratios (across all models) in the NCDS cohort were also larger than in the BCS70 cohort. Further, after considering the role of adult predictors, more robust associations between early life domains and hypertension and obesity comorbidity remained in the NCDS cohort compared to the BCS70 cohort. These three factors may indicate that obesity-hypertension comorbidity is more closely related to the domain-specific risk scores in the NCDS compared to the BCS70.

For the BCS70 cohort, the parental and family environment domain was a robust predictor of obesity-hypertension, but in the NCDS cohort, it was not a predictor. One explanation for this difference is the wider demographic and social context of the parental-family environment in which each cohort grew up. For the NCDS cohort (born 1958), most children grew up in households dominated by a stable nuclear-family model [37]. However, the BCS70 cohort (born 1970) grew up during a period of rapid change within the parental-family environment. The liberalisation of divorce laws coupled with changing attitudes and norms relating to marriage were reflected in a sharp rise in divorce rates and a decline in first marriage rates from the 1970s onwards [38,39]. The BCS70 cohort grew up during a period of increasing rates of parental partnership dissolution, single parent households and a rise in children living with non-biological parents and non-full siblings, and this is reflected by the retained variables following stepwise backwards elimination (step 1 of the analysis). Specifically the BCS70, unlike the NCDS, retained variables relating to the father of the cohort member including father figure and if the father helps manage the cohort member. Therefore, it could be hypothesised that the impact of these rapid changes to the parental-family environment, particularly around the role of fathers, had a greater impact on the BCS70 cohort compared to the NCDS who were less exposed to these changes. This is demonstrated in a paper that found women in the BCS70 who experienced parental separation were more like to have hypertension at age 46, compared to those who had not experienced a parental separation in childhood [40].

A further cohort effect is within the antenatal, neonatal, prenatal and birth domain which was a robust predictor of obesity-hypertension comorbidity in the NCDS cohort but not in the BCS70 cohort. It is likely that even over this short 12-year period, obstetric, neonatal and speciality care for ill infants will have improved [41], thus potentially reducing the impact of antenatal, neonatal, prenatal and birth adversity in the younger cohort (BCS70). Another difference between the antenatal, neonatal, prenatal and birth domain was that the stepwise backwards selection (step 1 of the analysis) retained different variables. The variables retained in the NCDS cohort focused on the health of the child at birth whereas, for the BCS70 cohort, the retained variables focussed on factors relating to maternal fertility history and maternal health behaviours during pregnancy.

In the NCDS the prevalence of obesity, hypertension and combined hypertension-obesity was lower than observed in the BCS70 cohort [18], suggesting that those with the outcome in the NCDS cohort may be a more select group of individuals. However, given that blood pressure increases with age [42] and that the BCS70 cohort were slightly older than the NCDS cohort at outcome data collection, it is important to note this relationship between blood pressure and age might explain some of the increased prevalence of obesity-hypertension amongst the older BCS70 cohort. In the BCS70 study we additionally classified hypertension if a participant had received a doctor’s diagnosis of high blood pressure or hypertension at age 46 (self-reported) even if the blood pressure measurement was less than 140/90 mm Hg, to account for those who may have had a lower blood pressure reading due to hypertension medication. In the NCDS self-reported hypertension was not asked.

There are a number of important next steps. We have identified robust domains that predict obesity-hypertension comorbidity, the next step therefore could include modelling prevention scenarios within these domains to better inform policy to help people live in better health for longer. Secondly, it is important to expand the methods presented here to consider the relationship to other multimorbidity clusters and outcomes that develop a more sophisticated understanding of multimorbidity, including focussing on burdensomeness and complexity for example incorporating eight themes of ‘work’ burden for those living with multimorbidity [43].

### Strengths and limitations

The NCDS dataset allowed us to explore a wide array of social, environmental, and family variables in childhood to represent five early-life domains. The data also afforded the opportunity to analyse potential adult predictors and objective measures of both obesity and hypertension measured at midlife.

A limitation of the NCDS cohort was the significant under-representation of individuals from non-white ethnic backgrounds and therefore the cohort does not reflect the ethnic diversity of the British population today. Further using BMI (to indicate obesity) is limited given the established research that has demonstrated the measure risks overestimating body fat in those who have muscular builds [44]. Another limitation of the hypertension outcome was that we were unable to consider those who were on antihypertensive medication, which could have lowered blood pressure readings at the time of cohort measurement, and these individuals could have been mis-counted in the non-outcome group.

Given our methods explored domains that were derived from combined risk factors, as opposed to considering these risk factors individually, there is a possibility that our prediction models are over-adjusted [45]. However, we felt this was unavoidable given our research question was to consider domains rather than individual risk factors. Further, as we have argued previously, we know that children’s early life experiences are intersecting, and therefore research must explore methods that can analyse multiple domains simultaneously, as this represents the best approach to disentangle and understand the role of competing early life domains for future health outcomes. In previous research, we considered alternative methods to achieve the aims of this paper such as deriving each variable (within each domain) into a binary (yes/no) outcome, and then summing to produce a count of adversity within each domain [46]. However, this approach was more limited (than the one presented here), given we were required to assume that all variables (within each domain), and all domain risk scores, carried equal weight. Secondly, by deriving variables into a binary indicator we disregarded information contained within the original data structure and for most variables we had to implement arbitrary binary cut-off points.

### Conclusions

We have demonstrated there are early life course domains that are robust predictors of obesity-hypertension comorbidity across two longitudinal cohorts. There were some differences across cohorts for the antenatal, neonatal, prenatal and birth domain and parental and family environment domain; we suggest these differences could be due to a cohort effect of the environment in which each cohort grew up. Overall, shared early life characteristics could have a role in predicting obesity-hypertension comorbidity, particularly for those who faced socioeconomic disadvantage in early life. These findings strengthen the case for ensuring public health programmes aimed at giving children the best start in life continue to be supported.

## Author Contributions

S.F., N.A., R.H., S.P., R.O., S.S. and A.B. contributed to the conceptualisation of the MELD-B project. S.S., N.A., and S.F. obtained the datasets. All authors contributed to the conceptualisation of the paper. S.S., and N.A. led the design and planning of the paper. R.O. led the design of the statistical analysis. S.S., N.A., A.B., N.Z., R.H., and R.O. supported the design, planning and reviewing of the statistical analysis. S.S. performed the statistical analysis with support from N.Z. S.S. prepared all figures and graphs. S.S., and N.A. produced the initial draft of the manuscript. All authors were involved in editing and reviewing the manuscript, and approved the final manuscript. S.S., N.A., and S.F. take responsibility for the data and research governance.

## Data Availability Statement

The NCDS datasets generated and analysed in the current study are available from the UK Data Archive repository (available here: http://www.cls.ioe.ac.uk/page.aspx?&sitesectionid=795).

## Acknowledgement

We would like to acknowledge all other members of the MELD-B Consortium, and we thank the participants of the NCDS cohort studies.

