## Supplementary Materials for "Early Life Predictors of Obesity and Hypertension Comorbidity at Midlife: Findings from the 1958 National Child Development Study (NCDS)"

Prenatal, antenatal, neonatal and birth domain  (variables collected at birth)

Maternal age at time of cohort member’s birth

Number of antenatal visits

Parity

Labour duration

Foetal distress

Birth interval

Pregnancy abnormality

Child resuscitated at birth

Child admitted to hospital at birth

Baby illness at birth

Developmental attributes and behaviour domain (variables collected at age 11)

Walking backwards in a straight line

Stand on right foot

Stand on left foot

Standing heel to toe

Hand control

Coordination

Inconsequential behaviour^1^

Nervous symptoms^1^

Socioeconomic factors domain (variables collected at age 11)

Tenure of accommodation

Child shares bedroom

Access to household amenities

Person per room

Father social class

Financial hardship

Free school meals

Child education and academic ability domain (variables collected at age 11)

Child’s general knowledge – teachers assessment

Child’s number ability – teachers assessment

Child’s oral ability – teachers assessment

Child uses books – teachers assessment

General ability test score^2^

Maths test score

Parental and family environment domain (variables collected at age 11)

Child’s mother figure

Child’s father figure

Family walks

Father supports with managing child

Mother interested in child’s education

Father interested in child’s education

*Supplementary Materials Table 1. All variables that were initially considered for selection within each domain*

*^1^These measures are taken from the Bristol Social Adjustment Guide (BSAG)*

*^2^Measure of general ability, including verbal and non-verbal elements*

*Supplementary Materials Table 2. Pearson’s Correlation Matrix. Multiple Imputation.*

|  | R Child education and academic ability domain predicted probability | Developmental attributes and behaviour domain predicted probability | Prenatal, antenatal, neonatal and birth domain predicted probability | Parental and family environment domain predicted probability | Socio-economic factors domain predicted probability |
| --- | --- | --- | --- | --- | --- |
| Child education and academic ability domain predicted probability | 1.00 |  |  |  |  |
| Developmental attributes and behaviour domain predicted probability | 0.10 | 1.00 |  |  |  |
| Prenatal, antenatal, neonatal and birth domain predicted probability | 0.00 | 0.04 | 1.00 |  |  |
| Parental and family environment domain predicted probability | 0.31 | 0.07 | -0.04 | 1.00 |  |
| Socioeconomic factors domain predicted probability | 0.25 | 0.05 | -0.01 | 0.29 | 1.00 |
| *Sample* | *9150* | | | | |

|  | | | Coef. | | [95% Conf | Interval] |
| --- | --- | --- | --- | --- | --- | --- |
| Resuscitation at birth: base no | | | 0 | | . | . |
| Yes | | | .489 | | -.096 | 1.075 |
| Hospital admission at birth: base no admission | | | 0 | | . | . |
| Admission for toxaemia | | | .58 | | .248 | .912 |
| Admission for other condition(s) | | | .128 | | -.192 | .448 |
| Baby illness at birth: no illness | | | 0 | | . | . |
| Illness | | | .451 | | -.074 | .976 |
| Constant | | | -3.197 | | -3.316 | -3.078 |
| Imputations | 50 | Number of obs | | 9150 | | |
| Prob > F | 0.0007 |  | |  | | |
| F( 4, 30028.8) | 4.84 |  | |  | | |

*Supplementary Materials Table 3. Regression coefficients of the stepwise backward elimination model of the relationship between the antenatal, neonatal and birth domain variables with obesity-hypertension comorbidity. Multiple imputed data (50 Imputations).*

*Supplementary Materials Table 4. Regression coefficients of the stepwise backward elimination model of the relationship between the developmental attributes and behaviour domain variables with obesity-hypertension comorbidity. Multiple imputed data (50 Imputations).*

|  | | | Coef. | [95% Conf | | Interval] |
| --- | --- | --- | --- | --- | --- | --- |
| Poor physical coordination: base certainly applies | | | 0 | . | | . |
| Somewhat | | | -.239 | -.956 | | .478 |
| Not at all | | | -.421 | -1.071 | | .229 |
| Standing heel to toe: base very steady | | | 0 | . | | . |
| Slight. unsteady | | | .273 | .042 | | .505 |
| Very unsteady | | | .448 | .088 | | .807 |
| Constant | | | -2.867 | -3.519 | | -2.214 |
| Imputations | 50 | Number of obs | | | 9150 | |
| Prob > F | 0.0001 |  | | | | |
| F( 5, 19136.8) | 5.45 |  | | | | |

*Supplementary Materials Table 5. Regression coefficients of the stepwise backward elimination model of the relationship between the socioeconomic factor domain variables with obesity-hypertension comorbidity. Multiple imputed data (50 Imputations).*

|  | Coef. | | [95% Conf | | Interval] | |
| --- | --- | --- | --- | --- | --- | --- |
| Father social class: base Social Class I | 0 | | . | | . | |
| Social class II | .061 | | -.612 | | .734 | |
| SC III non-manual. | .45 | | -.254 | | 1.154 | |
| SC III manual | .791 | | .181 | | 1.401 | |
| SC IV non-manual | 1.287 | | .476 | | 2.098 | |
| SC IV manual | .768 | | .121 | | 1.414 | |
| Social class V | 1.272 | | .584 | | 1.959 | |
| Constant | -3.758 | | -4.345 | | -3.171 | |
| Imputations | | 50 | | Number of obs | | 9150 |
| Prob > F | | 0.0000 | |  | |  |
| F( 6, 12650.7) | | 6.07 | |  | |  |

*Supplementary Materials Table 6. Regression coefficients of the stepwise backward elimination model of the relationship between the education and academic ability domain variables with obesity-hypertension comorbidity. Multiple imputed data (50 Imputations).*

|  | | | Coef. | [95% Conf | Interval] | |
| --- | --- | --- | --- | --- | --- | --- |
| General ability test | | | -.015 | -.024 | -.005 | |
| Child oral ability: base exceptional | | | 0 | . | . | |
| Above average | | | .442 | .089 | .795 | |
| Average | | | .678 | .202 | 1.155 | |
| Child's gen knowledge: base exceptional | | | 0 | . | . | |
| Above average | | | -.149 | -.474 | .176 | |
| Average | | | -.494 | -.938 | -.05 | |
| Constant | | | -2.659 | -3.238 | -2.081 | |
| Imputations | 50 | Number of obs | | | | 9150 |
| Prob > F | 0.0001 |  | | | |  |
| F( 5, 19136.8) | 5.45 |  | | | |  |

*Supplementary Materials Table 7. Regression coefficients of the stepwise backward elimination model of the relationship between parental and family environment domain variables with obesity-hypertension comorbidity. Multiple imputed data (50 Imputations).*

|  | | Coef. | | [95% Conf | | Interval] |
| --- | --- | --- | --- | --- | --- | --- |
| Fathers interest in education: base very interested | | 0 | | . | | . |
| Some interest | | .261 | | .008 | | .515 |
| Little interest | | .314 | | -.03 | | .659 |
| Can’t say/no father | | .25 | | -.117 | | .618 |
| Family walks: base most weeks | | 0 | | . | | . |
| Occasionally | | .189 | | -.037 | | .416 |
| Hardly ever | | .43 | | -.003 | | .864 |
| Constant | | -3.371 | | -3.572 | | -3.169 |
| Imputations | 50 | | Number of obs | | 9150 | |
| Prob > F | 0.0253 | |  | |  | |
| F( 5, 14196.6) | 2.56 | |  | |  | |
